# Beekeeping for Mental Health Prevention and Wellbeing: A Pro-Environmental Qualitative Study in an Underserved Adolescent Community

**DOI:** 10.1101/2025.09.03.25334765

**Authors:** Sena Demir-Kassem, Rob Deeks, Ciara McCabe

**Affiliations:** School of Psychology and Clinical Language Sciences, University of Reading, Reading, United Kingdom; Together As One, 29 Church Street, Slough SL1 1PL, United Kingdom

**Author notes:** Correspondence Prof Ciara McCabe (PhD, FHEA), School of Psychology and Clinical Language Sciences, University of Reading, Reading, RG6 7BE, United Kingdom. Beekeeping and Mental Health in Young People.

## Abstract

**Background:** Untreated mental health difficulties in adolescence increase the risk of poor outcomes in adulthood. Preventive interventions can improve wellbeing but remain underutilized, particularly among underserved youth who experience high stress and limited access to support. This study examined whether engaging in a pro-environmental activity, beekeeping, could promote mental health and preventive wellbeing benefits in adolescents.

**Methods:** Sixteen adolescents (aged 14 to 18) from the youth-led charity Together As One, Slough, UK, which supports vulnerable young people from low-income and marginalized backgrounds, participated in a 12-session beekeeping program. Sessions combined hands-on hive management, educational workshops, and apiary visits. Program impact was explored through semi-structured interviews and a focus group, with data analysed thematically.

**Results:** Four themes were identified: (1) Motivations and challenges: curiosity, social encouragement, occasional fear and boredom; (2) Psychosocial benefits: reduced stress, enhanced emotional wellbeing, greater energy and structure, and stronger social connections; (3) Reconnecting with meaning: engaging in new activities, stepping outside comfort zones, and overcoming symptoms such as anhedonia; and (4) Environmental awareness: increased understanding of ecological systems and sustainability.

**Conclusion:** Beekeeping shows promise as a community-based, pro-environmental activity that can foster stress reduction, emotional regulation, social connectedness, and a sense of purpose in adolescents. These findings suggest that youth-led, nature-based interventions may represent an innovative and underutilized approach to preventive mental health support in underserved populations.

## Introduction

A recent study reports a global prevalence of depression in children and adolescents at approximately 33%, significantly affecting their physical health, mental health, and academic performance (Mei & Wang, 2024). Yet only 38% of affected youth receive treatment, and this number drops drastically to just 6% in lower-income regions (Wang et al., 2023). Untreated mental health problems in adolescence predict worse outcomes for mental health in adulthood (Otto et al., 2021). Preventive interventions have the potential to significantly improve mental health outcomes but remain underutilized (Singh et al., 2022).

According to Patel et al. (2023), a vital step toward addressing this issue is shifting from an over-reliance on industrialized treatment models to community-based mental health initiatives. These initiatives prioritize youth involvement in decision-making and amplify their voices, ensuring that mental health services are truly youth-focused, accessible, and effective. (Patel et al., 2023; Smith et al., 2024). Beyond participation, young people should also be actively involved in decision-making regarding the types of projects available in their communities. Youth Participatory Action Research (YPAR) has emerged as an effective strategy to amplify young voices, ensuring that mental health interventions are youth-centered and evidence-based (Smith et al., 2024; Watson et al., 2023). Engaging young people not only improves the quality, relevance, and impact of research but also fosters personal growth, empowerment, and skill development among participants (Watson et al., 2023).

Adolescence is a particularly stressful period marked by rapid developmental changes and increased vulnerability to depression, anxiety, and external stressors(Romeo, 2013; Romeo, 2017). Adolescents may struggle to make decisions that support their mental health and are more prone to impulsive behaviours, which can be worsened by daily stressors that impair decision-making (Do et al., 2022). Building resilience is essential to buffer against these negative effects (Anyan & Hjemdal, 2016).

One of the most effective ways to foster resilience is through structured activities and hobbies that promote well-being. Participation in such activities enhances social connections, encourages personal growth, and fosters skill development, all of which contribute to mental resilience (Bälter et al., 2023; Durlak et al., 2010; Kusier et al., 2024; Ramey & RoselJKrasnor, 2012; Trainor et al., 2010). Community-based initiatives play a key role in offering these opportunities, providing structured recreation that supports young people (Bälter et al., 2023; Kusier et al., 2024; Torres Sanchez et al., 2022). As many communities face challenges related to isolation and loneliness, which can negatively affect well-being (Hall et al., 2024), programs that build social and emotional skills are particularly valuable (Bälter et al., 2023; Durlak et al., 2010; García-Poole et al., 2019; Kusier et al., 2024). It is especially important that such initiatives reach underserved and underrepresented youth, who experience higher stress and reduced access to mental health resources (Iwasaki & Hopper, 2017; Kusier et al., 2024; Schaffalitzky et al., 2015).

A wide variety of recreational activities are offered through these projects, including workshops, cooking, sports, animal interaction, nature excursions, creative arts (Bälter et al., 2023; García-Poole et al., 2019; Kusier et al., 2024). Nature-based activities, in particular, are increasingly recognized for supporting mental health (Lackey et al., 2021; Zamora et al., 2021), buffering against the negative effects of high screen time for example (Oswald et al., 2020). A study found that adolescents who spent more time in nature during the COVID-19 lockdown reported fewer declines in well-being (Jackson et al., 2021). Beekeeping, in particular, is one such activity, regular engagement in beekeeping has been reported to improve mental health by reducing stress, enhancing positive emotions, and strengthening connections with both nature and others (Burke & Corrigan, 2024; Whitaker, 2022). While many studies focus on experienced beekeepers, beekeeping has also shown therapeutic benefits for young people, including stress relief, increased happiness, and improved well-being (Ciraulo et al., 2024).

The connection between mental health and meaning in life further supports the value of beekeeping as it offers an opportunity to integrate meaning with action, providing participants with a purposeful, structured, meaningful experience (Alton & Ratnieks, 2022). Previous research has shown that a sense of meaning is closely linked to reduced depression symptoms (Demir-Kassem et al., 2025), while other studies indicate that engaging in meaningful activities can enhance mood and alleviate depression (Crego et al., 2021; Nagata & Kono, 2022).

Simultaneously, concern over climate change is increasingly prevalent among young people, contributing to heightened psychological strain (Ojala et al., 2021; Pihkala, 2020; Stewart, 2021). For some, the belief in inevitable environmental doom undermines their ability to engage with a sense of meaning (Guthrie, 2023). However, pro-environmental activities such as beekeeping can counteract these effects by improving well-being and offering a sense of agency (Capaldi et al., 2015). By fostering a connection with nature and promoting prosocial behaviours (Pirchio et al., 2021), beekeeping provides an avenue for addressing eco-anxiety while simultaneously enhancing mental health and well-being.

### 1.1. Youth-Led Beekeeping Initiative

We report on a community-led pilot in which the youth charity *Together as One* (TAO), supported by Community Volunteer Services, developed and delivered a beekeeping project to examine its effects on youth mental health. The research formed part of the British Science Association’s Community Engagement work, funded by UK Research and Innovation (UKRI), and was co-created with the BSA, the authors’ university, and underrepresented communities (see Supplementary Doc).

### 1.2. The aims of the project

The aim of the beekeeping project was to help young people connect with others, develop skills, practice mindfulness, and be more active to support their mental health and wellbeing. We used semi-structured interviews and a focus group to explore the benefits of pro-environmental behaviours, as engaging with nature and sustainability is linked to reduced stress and improved mental health, including depression and anhedonia (Capaldi et al., 2015; Ciraulo et al., 2024; Lackey et al., 2021; Watson, Harvey, et al., 2021; Zamora et al., 2021). We also examined meaning in life, given its association with lower depression and anhedonia in youth (Demir-Kassem et al., 2025) and explored participants’ views on improving youth mental health initiatives.

## Method

We followed the Coreq checklist and reported the study details accordingly (Tong et al., 2007).

### Participants

Sixteen adolescents aged 14–18 yrs. participated in the beekeeping project led by TAO. The group included individuals with additional support needs, such as learning or physical disabilities, and mental health concerns including depression and anxiety.

Young people aged 16 and over completed an online consent form directly. For those under 16, parental or guardian consent was obtained first, followed by the young person’s online assent.

### Ethical Considerations

Procedures complied with national ethics standards and the Helsinki Declaration, approved by the University Ethics Committee (Ref: 2024-009-CM). Participants were informed of the study’s purpose, withdrawal rights, and data confidentiality. Pseudonyms and secure, GDPR-compliant storage protected identities, and authors completed DBS checks for safeguarding

### Overall design

This study employed an exploratory qualitative design without a predetermined theoretical framework, allowing themes to emerge inductively from participant narratives. Semi-structured interviews and a group discussion were used to explore the effects of the beekeeping project, enabling an in-depth understanding of participants’ experiences and perceptions.

### Beekeeping Procedure

Participants, aged 14-28 yrs, completed 12 structured sessions combining theoretical instruction and practical experience (Table 1) between Feb and Mar 2024.

**Table 1.** The Details of Youth Beekeeping Program: Session Topics & Schedule.

|  |  |  |
| --- | --- | --- |
| February 7<sup>th</sup> 2024 | The Honeybee | - Bees, Wasps & Hornets |
|  |  | - Workers, Drones, and Queens<br>- Anatomy and life cycles<br>- Wax production, comb building |
| February 21<sup>st</sup> 2024 | The Bee Hive | - How Beehives work<br>- How to construct them<br>- Supers, feeders, excluders, etc.<br>- Possibly Frame preparation |
| February 28<sup>th</sup> 2024 | The Beekeeping Year | - The beekeeping year and what to do<br>- The four seasons in detail |
| March 13<sup>th</sup> 2024 | The Queen | - Life cycle<br>- Supersedure<br>- Swarming<br>- Swarm management |
| March 20<sup>th</sup> 2024 | Disease and Pests | - Identifying "Healthy"<br>- Varroa, other diseases, and treatment<br>- Pests, predation, robbing, and prevention |
| March 27<sup>th</sup> 2024 | Getting Started | - Building your Apiary<br>- Bee suits<br>- CBS trading |
|  |  | - Hive Products |
| April-May 2024 | 3 X Apiary Visits | -Observing the colonies<br>-Hive inspection<br>-Identifying the queen |
| June 29<sup>th</sup> 2024 | Follow-up Hive Assembly | -Finalizing hive structures if any were unfinished.<br>-Preparing for mid-season honey storage and pest control. |
| September 18<sup>th</sup> 2024 | Pest Control & Hive Health | -Counting mites to assess infestation levels.<br>-Potential treatments if needed. |
|  | Online activity | -Young people vote on names for new queens. |
| Summer 2025 | Harvesting | -Extracting honey.<br>-Jarring and donating honey to Slough Food Bank. |

### Questionnaires

*Mood and Feelings Questionnaire* (MFQ) (Costello & Angold, 1988) is a widely used measure of depressive symptoms and is suitable for adolescents (Burleson Daviss et al., 2006; Jarbin et al., 2020). 33 items scored on a three-point scale, with a maximum score of 66. Higher scores reflect greater depressive symptoms; scores above 10 indicate moderate depression, and scores over 27 indicate clinical levels. MFQ shows excellent internal reliability, with Cronbach’s alpha above .90.

*Anhedonia Scale for Adolescents* (ASA) (Watson, McCabe, et al., 2021), is a 14-item measure specifically designed for adolescents, with scores ranging from 0 to 42. Scores of 16 or higher indicate clinical levels. ASA captures multiple components of anhedonic experience and demonstrates excellent internal reliability (Cronbach’s alpha = .94)

*Meaning in Life Questionnaire* (MLQ) (Steger et al., 2006) assesses both the perceived presence of and active search for meaning in life. It is widely used and considered suitable for adolescents (Rose et al., 2017). The scale includes 10 items rated on a 7-point Likert scale, with total scores ranging from 0 to 60. Higher scores reflect a stronger sense of meaning. MLQ shows strong internal consistency, with Cronbach’s alpha ranging from .82 to .88.

### Interviews

The semi-structured interviews and group discussion were conducted by the first author.

In the final weeks of the program, participants were invited to take part in interviews. Seven (P1, P2, P3, P4, P5, P6, P7) completed individual interviews of approximately 20 minutes via Microsoft Teams, and five (P4, P5, P6, P8, P9) joined a one-hour group discussion. Both formats used open-ended questions to explore participants’ experiences in the project, perceived effects on well-being, environmental awareness, and meaning in life. Questions are provided in the supplementary doc. Sessions were recorded and automatically transcribed by Microsoft Teams. Transcripts were cross-checked against audio recordings by the researcher for accuracy, and efforts were made to ensure verbatim transcription. No field notes were taken. All interviews occurred in private to ensure comfort and confidentiality. However, three community researchers and team leader were present during the group discussion.

The primary researcher’s academic background in depression, anhedonia, meaning in life, and pro-environmental behaviour informed the study’s focus. These interests shaped the questions, especially those examining how beekeeping might support mood, motivation, enjoyment, and existential reflection.

### Data analyses

Data were analysed using thematic analysis, following Braun and Clarke’s six-phase framework(Braun & Clarke, 2006; Byrne, 2022). (1) Familiarization with data: transcripts were read multiple times to develop a thorough understanding. (2) Generating initial codes: relevant segments were coded systematically across the dataset. (3) Searching for themes: codes were grouped into potential themes reflecting recurring patterns. (4) Reviewing themes: themes were refined to ensure they accurately captured the data and aligned with the study’s aims. (5) Defining and naming themes: each theme was clearly articulated to represent its core concept. (6) Producing the report: themes were situated within the broader literature on adolescent mental health, meaning in life, and environmental awareness. Coding was conducted by the primary researcher in collaboration with the senior author, following Braun and Clarke’s (2006) framework. NVivo 14 software was used to organize and manage the data. To ensure accuracy and trustworthiness, transcripts were analysed verbatim, and direct participant quotes were used to support the identified themes (see Results and Supplementary doc).

## Results

Participants had a range of depression, anhedonia and meaning in life scores, 50% of participants had moderate to high depression and anhedonia symptoms (Table 2).

**Table 2.** Descriptive statistics and demographic characteristics of the participants.

|  | Gender | Ethnicity | *MFQ* | *ASA* | *MLQ* |
| --- | --- | --- | --- | --- | --- |
| Individual Interviews |  |  |  |  |  |
| P1 | Female | White: English | 13 | 9 | 27 |
| P2 | Male | Asian / Asian British: Indian | 2 | 6 | 25 |
| P3 | Male | White: English | 6 | 5 | 26 |
| P4 | Male | White: British | 26 | 17 | 8 |
| P5 | Female | Black African | 18 | 9 | 51 |
| P8 | Female | Mixed: Black African and Black Caribbean | 16 | 12 | 26 |
| P9 | Male | Black Caribbean | 7 | 7 | 27 |
| Focus group |  |  |  |  |  |
| P4 | Male | White: British | 26 | 17 | 8 |
| P6 | Female | Black African | 18 | 9 | 51 |
| P7 | Female | Mixed: White and Black Caribbean | 33 | 19 | 30 |
| P10 | Female | Mixed: White and Black Caribbean | 10 | 14 | 35 |
| P11 | Female | Mixed: Black African and Black Caribbean | 16 | 12 | 26 |

Abbreviations: *MFQ*; Mood and Feeling Questionnaire, *ASA*; Anhedonia Scale for Adolescents, *MLQ*; Meaning in Life Questionnaire

Four themes emerged: Motivations, Psychosocial Benefits, Reconnecting with Meaning, and Environmental Awareness (Figure 1).

**Figure 1:**
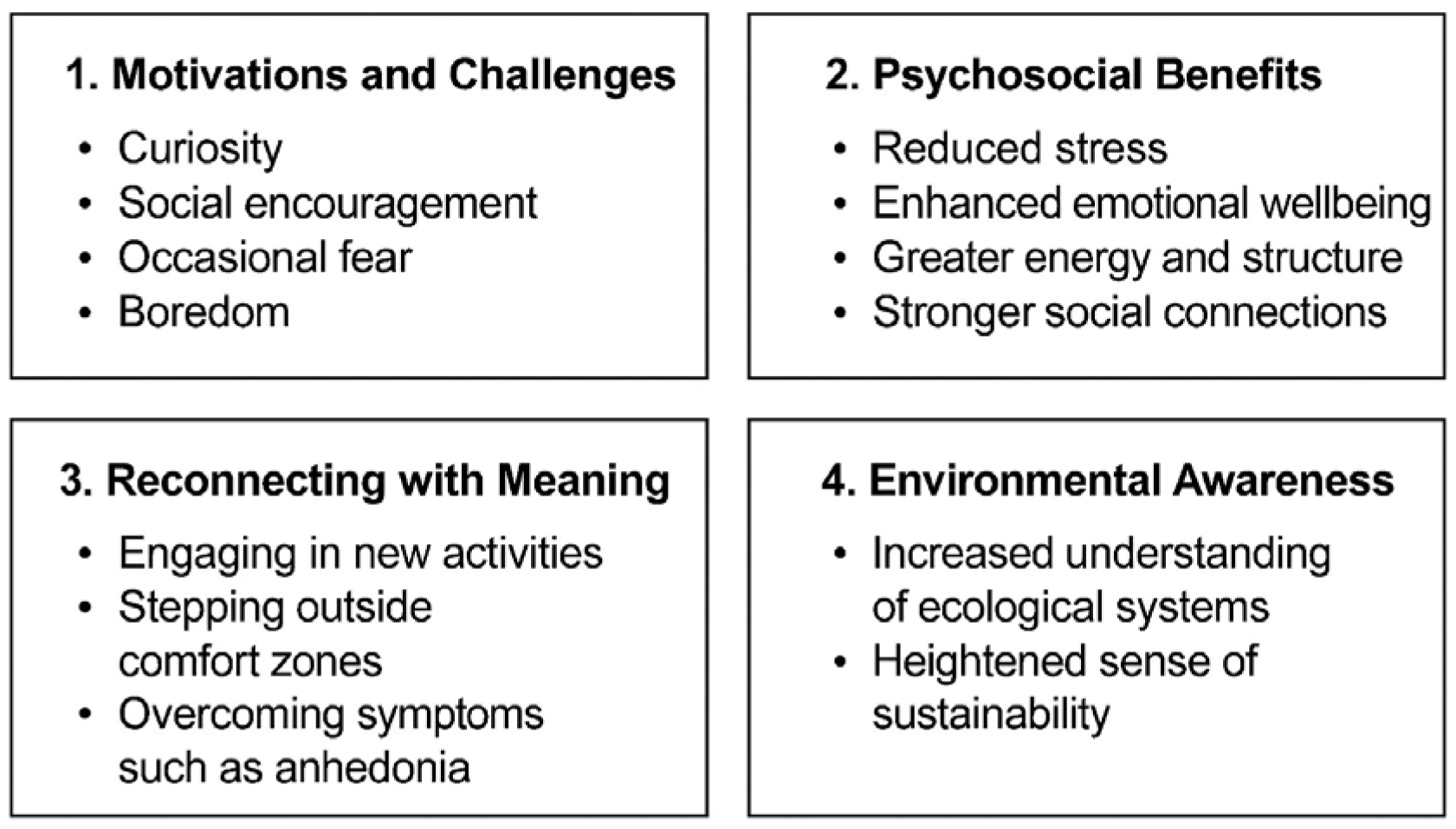
Themes and sub-themes related to beekeeping project

## Theme 1: Motivations and Challenges for Engagement

### Subtheme 1.1: Motivations

The participants’ initial reactions to the beekeeping project were diverse, ranging from enthusiasm and curiosity to neutrality and external influences like parental encouragement.

For example, P1 shared their motivations, despite uncertainty about the project’s details.

> I was excited because I love animals, but I didn’t really know what it entailed very much, but I was excited (P1).

Others stating their decision to participate being shaped by parental encouragement.

> I think it’s good.…. but my mom really signed me up to it. I don’t really stand up for myself that much because I’m not really…on the beekeeping (P2).

There was an appreciation for the potential benefits the project offered, particularly the opportunity to develop new skills or hobbies.

> Well, it’s good to learn a new skill. Learning how to do beekeeping is interesting… Maybe you know, discover a hobby, side hobby which I think I might have (P3).

Others were primarily motivated by social aspects.

> I didn’t really know much about beekeeping. I saw it more as like a social opportunity to be with my friends but at the same time I was also quite interested in beekeeping because obviously it’s beekeeping like no one really talks that much about it (P5).

### Subtheme 1.2: Challenges

While the project was largely positive, participants noted some challenges. Theory-based sessions were described as “boring,” “not engaging enough,” or “like school,” in contrast to the excitement of practical activities.

> When they have practical aspects, when we’ve brought out bees, and the different types of frames, that was interesting, but then when it was just like one PowerPoint but for 2 hours, that was a bit boring (P6).

However, some said they enjoyed both and appreciated the knowledge they gained “Obviously very different settings, but I think both settings were great, and the people were lovely and I learned a lot”. (P1).

Fear and apprehension about bees were common challenges P8 acknowledged her initial fear of being stung, “we went to see our hive and then I stood in like a bee suit. And we opened the hive for the first time and all my fears were gone because I knew what bees want.” Over time, participants overcame their fears, crediting the supportive environment and knowledge about bees for building their confidence.

## Theme 2: Psychosocial Benefits

Consisting of three subthemes: (1) improving emotional well-being and providing stress relief, (2) enhancing routine and energy levels, and (3) building social connections and support. While these aspects were overwhelmingly positive, participants also highlighted some (4) challenges and negative aspects.

### Subtheme 2.1: Improving Emotional Well-being and Providing Stress Relief

Participants either expressed directly or hinted that their mood had lifted and that they felt a heightened sense of happiness be being in the project. For many, it served as a source of pleasure and something to anticipate with enthusiasm.

> it really boosts my mood to be around them, do something about, being around animals…..I think all of it helped, it boosted my mood, the information sessions, I found them very interesting and a proper boost in my mood (P1).

> During the project, I think that was my peak happiness. I was really happy. I was enjoying my time (P5).

Participants consistently described how the sessions became a welcome break from the pressures of school and daily routines. During the project they said joining beekeeping sessions was a stress-relieving activity and an uplifting break from their routines.

> And it was a nice wind down from school…… It’s something to look forward to during the week…it’s more about the stress relief, (P5).

### Subtheme 2.2: Enhancing Routine and Energy levels

Participants highlighted the regularity of the project as a key benefit, bringing structure and energy to their weeks. The sessions became a source of anticipation, helping them develop a routine that felt enjoyable rather than obligatory. P8 shared, “it was nice to get out of the house and go to do something that’s not in my normal routine.” Also the novelty of the project refreshed their schedules. P1 said “it definitely didn’t feel like school —more of a hobby.” P4 commented on its ideal frequency, saying “because I feel they’re not too often, so I kind of want to take part because I don’t get the enjoyment of having them on a really regular basis.”

The project helped participants develop a more open-minded perspective, maintain higher energy levels, and explore new hobbies and activities. P8 said “I was more interested in the world around me. And I had a more open mindset”.

### Subtheme 2.3: Building Social Connections and Support

Socialization was a prominent benefit of the project. Adolescents described how the sessions provided an opportunity to strengthen friendships and connect with new peers in a supportive environment. P6 said it was “a way to socialize with lots of people”. Although most of them knew each other, it was an opportunity to get to know them better.

> I already knew some of the people in the beekeeping group, but I didn’t really know that much about them.,,,I pretty much just knew their name but during the beekeeping group, I found out more about them (P8).

The project also enhanced their social skills and allowed them to meet new people.

> It made it easier to speak to people and be more confident to speak to people instead of just enclosing myself off to people I already knew…I find public speaking easier…socializing easier and I have lots of new friends now (P8).

The sessions fostered a sense of community and mutual support. P9 gives an example of support they felt with challenges they experienced during the project: “they helped me calm myself down and at the end of it I was able to pick it up (honeycombs)”.

## Theme 3: Reconnecting with Meaning

This theme captures broader suggestions for improving meaning in life and overcoming depression/anhedonia for adolescent mental health. They discussed common challenges and proposed ways to improve well-being through exploring new activities and building social connections.

### Subtheme 3.1: Foundations of a Meaningful Life

Most shared ideas about what might bring meaning, such as “living your life to the fullest”, “caring for people and getting cared for” and “family”, “helping others”, having “a purpose that you want to achieve”, and “just to be happy and to be content with what you’re doing”.

Having strong personal values was especially highlighted as an essential component for constructing a meaningful life. Participants emphasized that these values were vital for fostering authenticity and guiding one’s own path: “If you go into life with no core values…you’re just following what others are doing, and it’s not really your life you’re living.” (P6).

### Subtheme 3.2: Leaving the Comfort Zone and Exploring New Activities

Participants stressed the importance of stepping outside their comfort zones to discover meaning and improve emotional well-being. One participant specifically linked this with the need to stop being overly concerned about others’ opinions, suggesting that growth requires resisting social pressures: “*You should get out of your comfort zone…stop caring about what everyone thinks of you.” (P6)*.

Exploring new interests, despite initial discomfort, was considered beneficial, although many admitted that embracing this could be challenging:

> I feel you have to give yourself time to explore more hobbies…. I don’t really explore find out new hobbies …because I think I prefer to stay to something I know than going into something different. (P6).

Participants consistently advised peers to try various activities and encouraged an open-minded approach to discover what brings genuine enjoyment and meaning.

### Subtheme 3.3: Obstacles to Finding Meaning and Exploring Interests

These included fear of judgment or social embarrassment, financial concerns, reliance on parental support, and unease around unfamiliar social groups.

> Some people don’t explore other hobbies because they’re scared of embarrassment. Like some girls won’t join the football club because they’re scared of embarrassing themselves. (P11).

> But nowadays, if you’re new to something, a lot of people will judge you until you get used to that sport and used to people (P10).

Participants also pointed out that a lack of independence, both financially and in terms of mobility, often held them back from new activities, as they still needed parental support.

> if you want to try something new, like ice skating… you have to have a parent to drive you, to pay for it (P6).

> financial situations may also be an obstacle or even in a case where it could be you can afford to try something, but it’s the case of you’re trying to determine if you want to invest even anything to try it because you’re not initially sure on your enjoyment or aspiration to do it. (P4).

### Subtheme 3.4: Importance of Social Connections

Being around people was thought to play a big part in better well-being overall. P 11 noted that being alone can make it hard for some people to enjoy activities, saying: “I feel like some people have trouble to enjoy things because they’re alone most of the time.” Discussing your troubles with someone was seen as an effective way to ease distress: “I would talk to someone about it rather than just …let it build up…..let it get to me.” (P7). According to a participant, interacting with others also helped “it really helps me I’m, like, helping myself, but at the same time, I know I’m helping other people….I know that I’m doing a good deed for other people” (P10).

Some participants felt that staying socially active is important but acknowledged challenges and the need for limits. Issues like “not knowing anyone,” and dealing with dominant personalities in groups were seen as obstacles, sometimes making social situations isolating and stressful instead of enjoyable.

## Theme 4: Environmental Awareness

Three subthemes emerged: *Pro-environmental behaviours, Understanding the importance of bees*, and *Reflections on broader environmental issues*.

### Subtheme 4.1. Understanding the Importance of Bees

For participants, the project was a transformative journey. Initially, their knowledge about bees was limited to simple concepts, such as the “queen bee,” “little flying insects that live in strange looking colonies,” “eating honey,” and their role as “main pollinators.” Their awareness of bees’ significance was minimal, as P8 reflected, “I knew that they were important, but I just didn’t know in what way”.

Through hands-on activities, participants gained practical knowledge and a deeper appreciation of bees’ environmental impact.

> I know how important they are. I know their life cycles. I know how to spot diseases in the hives. I know how to take care of them now. It was very impactful….. it makes you really appreciate how much bees do for the environment. … they are the main pollinators and if we didn’t have bees, I think this is like a short time period where we would just run out of food because they are so impactful (P5).

> ..people have always said that bees have a huge impact on the world. But I never really believed them because I thought they used to just fly around and take all the pollen and stuff until, the beekeeping introduced me to actually what they’re doing in the world and made me realize would it be the same without bees? (P 10).

### Subtheme 4.2. Pro-environmental Behaviours

Most of the participants recognised “behaviours benefiting the environment”, “helping nature”, “being respectful to the environment”, “making a difference in the environment”. “To recycle”, “not litter”, “manage carbon footprint or carbon emissions” as pro environmental behaviours that could be “little but impactful” and “a good thing”.

> I look on a smaller scale, what you can do on a small scale…but when you do it enough it can actually have a big impact (P1).

Participants also admitted difficulties of adopting such behaviours. P2 stated, “It’s a good thing to an extent, it depends on what we have to sacrifice as well.” They particularly emphasized financial challenges, with P3 noting, “I know sometimes being ecofriendly can be a little bit more expensive.*”* They also underlined that many people are not able to do them.

> ..like most protesters do and so whilst it should be a good thing, it isn’t always necessarily shown in the greatest of ways (P4).

> wrappers, there’s some bits that can’t be recycled so… the bits that can be recycled are in there, and the bits that can’t aren’t, like small things like that, isn’t easy (P1).

### Subtheme 4.3. Broader Environmental Reflections

We explored whether the project encouraged participants to reflect on environmental issues, some reported more meaningful realizations.

For many participants, the project reinforced pre-existing ideas without fundamentally altering their perspective on the environment. One participant remarked, “I don’t think it really added anything to what I already think about, I already understood that bees were important to the environment” (P4). These reflections indicate that the project aligned with their existing beliefs.

For some participants, the project acted as a subtle reminder about bees, “It just made me think what important roles that the bees do and how they like impact us every day”. Another reflected “The climate change is affecting the bees, and the bees are important. So, it does make you think about climate change” (P3). These reflections show that the project prompted some thought about the environmental impact of bee conservation.

A few participants found the project made them think about systemic and societal responses to environmental issues and pointed to a perceived imbalance in how environmental responsibility is assigned. P11 expressed frustration saying,

> It’s a global thing and everyone has to do their part, and we can’t because what they do is they prey on us, and they guilt trip us … if you don’t recycle, you’re committing carbon footprints, or you don’t care about the planet… But at the same time, they’re letting people like our Prime Minister … go around the world to each vacation home in a private jet, which emits thousands of tons of carbon dioxide.

P 4 criticized brands for exploiting environmental concerns for profit, stating

> brands that promote being environmentally friendly do it for the value it gives them from people who want the face of doing their part to just gain more money. …no one really cares too much about the environment, because they understand it’s too far gone…..we just make money off it.

## Discussion

Youth-led community interventions are key to fostering resilience and preventing mental health issues, especially for underserved youth (Patel et al., 2023). This project explored how pro-environmental activities, specifically beekeeping, support adolescent wellbeing. Findings extend previous research (Burke & Corrigan, 2024; Ciraulo et al., 2024; Whitaker, 2022) across four themes: Motivations, Psychosocial Benefits, Reconnecting with Meaning, and Environmental Awareness.

### Motivations for engagement

Participants’ motivations for engaging in beekeeping were varied, spanning intrinsic curiosity, external encouragement, and the search for new experiences. While some expressed genuine enthusiasm, others joined at the suggestion of parents or peers, yet still reported enjoyment once involved. This reflects research showing that young people may struggle to initiate activities but often benefit once engaged (Watson, Harvey, et al., 2021) Badura et al. (2024). For some, the main motivation was social connection rather than beekeeping itself, highlighting the value of shared experience in adolescence (Sachser et al., 2018). Overall, engagement was shaped by a mix of curiosity, relational needs, and developmental opportunities, consistent with wider evidence on youth environmental action (Romano et al., 2024). These findings suggest that youth programs should balance skill-building with social connection and enjoyment to foster participation.

Although feedback was largely positive, some found theory sessions less engaging than hands-on work, likening them to school. Initial fears of handling bees were also common but eased with exposure and support. This highlights the value of participant feedback and adapting programs to sustain motivation, for example through shorter, more interactive or gamified sessions (Zhan et al., 2022).

### Psychosocial benefits

The beekeeping project provided adolescents with significant psychological and social benefits, including improved emotional well-being, daily structure, energy, and social connectedness. Participants described sessions as uplifting, offering a break from academic pressures and daily stress, consistent with research on the mental health impact of school environments (Kaczmarek & Trambacz-Oleszak, 2021; Murberg & Bru, 2004) (Moksnes et al., 2016; Ringdal et al., 2020). The program’s regularity fostered routine, anticipation, and engagement in meaningful activities, in line with existing literature on the positive effects of structured leisure activities (Bälter et al., 2023; Iwasaki & Hopper, 2017; Kusier et al., 2024; Trainor et al., 2010) contrasting with less beneficial unstructured leisure. Through hands-on beekeeping, young people developed tangible skills, experienced clear achievements, and built self-confidence. Many reported feeling more energetic, motivated, and open-minded, with positive effects extending beyond the project. These findings align with evidence that structured, adult-supervised activities promote skill development, goal attainment, and meaningful social interaction (Bälter et al., 2023; Kusier et al., 2024; Trainor et al., 2010).

Social connectedness was a key benefit, with participants reporting stronger relationships, improved communication confidence, and a greater sense of belonging (Kusier et al., 2024). As loneliness is linked to depression and anhedonia (Achterbergh et al., 2020; Prizeman & McCabe, 2025), youth-led community projects like this can help bridge gaps in social connection, potentially protecting against long-term physical and mental health effects (Park et al., 2020; Petitte et al., 2015; Wang et al., 2018).

### Reconnecting with meaning

Although young people in our study struggled to define ‘meaning in life,’ their insights reflected key components such as personal values, a unique life path, and a sense of purpose. While adolescents can discuss existential ideas as coherently as adults (De Vogler & Ebersole, 1983; Ratner et al., 2021; Shek, 2013) community projects could support reflection, value exploration, and goal-setting to help translate these insights into practice (Luz et al., 2017).

Participants highlighted the importance of engaging in enjoyable activities to combat low mood and anhedonia, though many struggle to identify what they enjoy (Watson et al., 2020). This may reflect a lower sense of self, linked to reduced meaning in life and greater anhedonic symptoms (Demir-Kassem et al., 2025). They suggested experimenting with new activities to discover enjoyment and better understand themselves. Community-led projects offer valuable opportunities for such exploration. Given that young people often spend much of their time alone despite enjoying social and leisure activities (Sahni & McCabe, 2025), creating accessible, engaging opportunities is increasingly important.

Young people reported barriers to exploration, including financial constraints, parental dependence, fear of judgment, and social anxiety. Limited access to nature and recreational activities, particularly for disadvantaged youth, can hinder well-being (Ghimire et al., 2014; Lovelock et al., 2016; Oncescu & Loewen, 2020; Waite et al., 2023). The beekeeping project highlights the mental health benefits of improving access to pro-environmental activities for underprivileged communities.

The young people also mentioned social anxiety as another challenge to engaging in new leisure activities alongside stigma and a fear of judgment. However, they described the beekeeping sessions as a more comfortable setting than school, where they could build social skills, form friendships, and feel accepted. This highlights the importance of creating inclusive, non-judgmental spaces where young people feel safe to explore new physical and social activities.

### Environmental awareness

Before the project, participants had only a basic awareness of bees, often limited to honey production or vague notions of pollination. The beekeeping sessions fostered a more nuanced understanding of bees’ ecological significance, particularly their role in sustaining biodiversity and ecosystem functioning (Patel et al., 2025). Such a shift in awareness, when combined with direct engagement in a nature-based activity like beekeeping, has the potential to influence future pro-environmental attitudes or behaviours (Krettenauer et al., 2024).

Most participants demonstrated a clear grasp of pro-environmental behaviour and believed their actions could make a difference. However, some hesitated to adopt such behaviours, perceiving them as personally costly or burdensome. This aligns with Prinzing (2023) who finds that environmentalism is frequently framed as a form of “self-sacrifice” something one must do “for the greater good” at personal cost. Although this kind of framing can create psychological distance and discourage engagement so we suggest environmental educational programs should frame environmentalism from the outset as personally rewarding. Our study supports this, providing evidence of the immediate and individual benefits of pro-environmental behaviours, such as improved well-being, autonomy, and quality of life (Prinzing, 2023).

A few participants expressed anger toward those they saw as primarily responsible for climate change, such as wealthy individuals, celebrities, and politicians, which raised questions about the fairness of individual responsibility. This kind of anger, when coupled with feelings of futility, may undermine motivation or lead to disengagement (Stanley et al., 2025).

Therefore, we propose future community-led environmental projects should emphasize to young people the benefits of collective action programs (Gulliver et al., 2022). How social capital (i.e., the community groups that facilitate social engagement) is key to the development of collective action (Putnam, 2000) and can produce shared norms of behaviour between community members and a generalized trust in individuals and institutions, facilitating civic action around societal issues (Wakefield et al., 2006).

A key strength of this project was its culturally diverse community. While stigma did not appear as a direct barrier, varying attitudes toward mental health may have influenced how openly participants discussed emotional struggles (Ahad et al., 2023) (Prizeman et al., 2024). Such cultural differences highlight subtle challenges for researchers in diverse settings.

## Conclusion

Community-based, experiential programs can boost youth mental health, especially for those with low mood or self-esteem, by fostering social connection and self-exploration. Sustained access for underserved youth acts as a protective, preventative measure. With rising mental health challenges, long-term funding and support for youth organizations and from governments is essential.

## Supporting information

Supplemental doc

## Data Availability

All data produced in the present study are available upon reasonable request to the authors

## Acknowledgements

We thank the young people of TAO for sharing their experiences, Slough Council for Voluntary Services for facilitating the project, Chalfonts Beekeepers for supporting the practical work, and the Jubilee Riverside Centre for hosting the hives. We are also grateful to Dr Sally Lloyd-Evans, Dr Alice Mpofu-Coles, and Kate Orchard (BSA) for their support in engaging local communities.

