## Supplemental doc for "Beekeeping for Mental Health Prevention and Wellbeing: A Pro-Environmental Qualitative Study in an Underserved Adolescent Community"

**Supplementary text**

**Extra quotes from themes**

**Theme 1: Motivations and Challenges for Engagement**

*Subtheme 1.2: Challenges*

Balancing the project with other commitments was a concern for one participant, who feared it might be overwhelming. As P4 expressed, “the amount of upkeep it’s gonna need.” However, P11 noted that the coordinator provided “a timetable for when we need to be outside of the house daily and so we'd know when to go, so it wouldn't be necessarily hard. It was just about following your time schedule, and the times didn't really change week to week.” This structured schedule helped make the project manageable.

*Subtheme 2.2: Enhancing Routine and Energy levels*

Without the project, participants felt fewer opportunities for meaningful activities after school. P6 admitted “I would just sleep if the project wasn’t scheduled that day. P1 added “it was really good to get out of the house......I think it got me out of my comfort zone a bit”. P4 and P7 highlighted that the activity added structure to their schedule, eliminating the need to decide how to use their time, which positively impacted their overall well-being.

*Having that regular basis where it is like every week on the same day helps, you think about how you have something going on rather than being unsure of what you're doing over the week. And I think that helps with the self-fulfilment part (P4).*

*routines help a lot of people to manage themselves easier. And just knowing that they have a certain thing on certain day, which is a lot easier for everyone (P7).*

P5 shared that her energy levels were significantly higher during the project than when she was solely focused on school, “during (the project) they were high because my body was getting used to like going out and studying for that long and after….now they I guess slumped”. The boost in energy levels and a more open mindset inspired participation in activities beyond school. P8 admits now “I'm more open to doing things rather than just like staying at home.” P4 said he started “doing some art activities” during the same period the project started.

*Subtheme 2.3: Building Social Connections and Support*

Participants emphasized how being part of the group reduced feelings of isolation and made them feel understood. The environment was inclusive and friendly, with familiar people, which made learning new things more accessible and less intimidating.

*Even though we were starting new, it was like a new hobby. You’re like comfortable with the people you were doing it with. So it was like the fear that you might have had doing something else, that because you didn't know who's there, that wasn't there because you know who's there (P6).*

They expressed that the supportive atmosphere during the project was missing in school, where they felt constrained and unable to be authentic. The lack of judgment during the project helped participants relax and engage more fully with the group.

*Like especially with this friendship that we have, you can just be yourself. People at school, they don't show their actual selves. They present themselves in a way that suits everyone that they hang with (P11).*

**Theme 3: Reconnecting with Meaning**

*Subtheme 3.1: Foundations of a Meaningful Life*

Participants expressed uncertainty about defining the meaning of life and admitted they hadn’t discovered their own. As P5 explained, “every time I think about it, I remind myself that I'm only, what, 15? So, I haven't seen the full excess of life.”

The importance of having a purpose in life was stressed by some of the young people, who saw it as key to providing direction and easing uncertainty. They believed that purpose helps focus on priorities, reduces stress, and brings clarity to future plans:

*If you have to set goals and a mindset for the future, it makes it easier because you know where to focus, what you should spend your time on* (P7).

Some participants felt that purpose was innate or something that gradually became clearer with time, "I think everyone has a purpose… It's just some people's purpose is easier to see, and some fall into their purpose later in life." (P5). One acknowledged the dual nature of purpose, seeing it as both inspiring and demanding, "It’s good in the sense that you have something you need to do, but also bad in the sense that you have something you need to do." (P4).

*Subtheme 3.4: Importance of Social Connections*

Participants underscored the importance of maintaining balance in social life.

*Being around people is fine, but you should also have time for yourself so that you can get to know yourself (P6).*

**Theme 4: Environmental Awareness**

*Subtheme 4.1. Understanding the Importance of Bees*

By the project’s end, participants gained not only knowledge but also a new perspective, shifting from fear or indifference to appreciation and awareness.

*As a kid…. I would just wish there were no bees because I really hated them. But now I realize how much the bees have an impact on the environment (P6).*

**Together as one**

TAO provides a wide range of services and activities for young people in Slough UK, and the surrounding areas. Founded in response to community tensions, TAO positions young people not just as beneficiaries but as key agents of change—making youth participatory action research a natural fit for its mission. While most of TAO’s opportunities are open access, the organization has become a trusted space for some of the community’s most vulnerable youth, offering both security and meaningful opportunities. As a result, TAO works extensively with young people from low-income backgrounds and those with adverse childhood experiences. In addition, TAO’s outreach—extending into settings such as the local Emergency Department—means they frequently support young people facing mental health challenges. Although TAO operates on the principle that everyone is welcome, the young people they serve often reflect a broad spectrum of vulnerabilities and complex needs.

TAO also plays an active role in various partnerships, including the Children and Young People’s Board and the Safer Slough Partnership. This involvement ensures a clear understanding of key issues—not only from daily interactions with young people but also through data and strategic insights from partners such as the Local Authority and the NHS. Recently, TAO previously collaborated with Rocket Science on a peer-led strategic needs assessment, in which young people conducted 191 interviews with their peers to explore their needs, concerns, and aspirations (RocketScience, 2025). Social isolation emerged as a significant issue, alongside a clear demand for more opportunities to bring different groups together. There was also a strong call for a more diverse range of mental health support, as well as greater access to positive youth activities. Therefore, there was a need for a project that simultaneously could address these priorities youth outlined.

On discussions of a potential partnership with the University on a youth-led project, young people were particularly passionate about environmental issues as they have a deep concern about climate change. The chance to engage in social action that directly benefits the planet was a key driver behind their vision for this initiative. This led TAO to organise a series of summer taster sessions, including fashion upcycling, bat conservation, litter picking, and beekeeping. The beekeeping session proved the most popular, and this formative experience inspired the young people to develop a vision for a young beekeepers’ project.

**Methodological reflections**

Several bee keeping societies were interested but had concerns over allergies, stings, and general risk involving children and young people. Despite widespread community recognition of bees’ importance, there was also a reluctance to accommodate hives near residential or commercial properties by landlords and local authorities. The first approved site was later withdrawn, and a second location required significant preparation, including the removal and recycling of scrap materials before installation could proceed. Transport logistics was another challenge. Parents and carers were understandably concerned about safety in the evenings, so the TAO team frequently arranged pick-up and drop-off of the young people, which lead to significant increases in time investment.

**Sustainability and long-term impact**

Ensuring the sustainability of community-led projects is critical to their long-term impact. In the case of this beekeeping initiative, sustainability is not just a goal but a necessity as caring for 45,000 bees means that the project will need to continue beyond the study period. This responsibility highlights the importance of maintaining young people’s engagement and leadership but also the need for continuous funding streams to allow the project to be continued and developed over time.

A. **Focus Group Questions**
1. How was your experience with the beekeeping project?
2. What aspects of the project did you enjoy or find boring?"
3. Do you think projects like this contribute to the wellbeing and mental health of youth? How?
4. Some young people feel a constant lack of motivation and can't enjoy their activities or life in general. Do you think it is common in youth? Why do you think this happens?
5. What kind of activities can help young people who are feeling depressed or experiencing anhedonia (explained as a lack of interest/motivation and joy in life)?
6. Do you believe being around other people can help with feelings of depression? In what ways?
7. Did this project make you think about the environment? Could you provide some examples?
8. Many people are concerned about environmental issues. Why do you think that is?
9. In your opinion, what are some effective ways to cope with worries about the environment?
10. Are you familiar with pro-environmental behaviours? What do they mean to you?
    a. Do you view them as positive or negative? Why?
11. What does having 'meaning in life' mean to you? Why do you think people feel unhappy when they find something to be meaningless?
12. How important is it to have purpose, goals, or values in life?
13. Can having a sense of purpose or meaning improve one's wellbeing? How?
14. How can young people find meaning or purpose in their lives? What actions should they consider starting or stopping?
B. **Individual interviews Questions**
1. What were your initial thoughts about the beekeeping project?
2. What was good about it?
3. Did you have any worries about it? If yes, what are they?
4. Does this project make you think about the worlds environment? Can you give examples
5. What were your expectations from participating in this bee project? How did it benefit you?
6. Are you familiar with the concept of pro-environmental behaviours? What does it mean to you?
7. How important do you think it is to take actions that benefit the environment?
8. How have you been feeling lately in general?
9. Have there been days when you felt really down, sad, or without much energy? How often does this happen?
10. Do you ever feel like you can’t be bothered? Or have low motivation, lack of interest in activities? Can you give me an example?
11. Can you tell me about some activities or hobbies you enjoy?
12. What does having a sense of purpose/ meaning in life mean to you? Can you give examples?
13. How important is it for you to find meaning or purpose in what you do? Is it good or a bad thing?
14. Can you describe how you usually spend your time? (with friends or in your community)
15. Are there activities involving others that you particularly enjoy?
16. How do you feel about working or doing activities with a group, like in the beekeeping project?
